# Predictive Machine Learning for Personalised Medicine in Major Depressive Disorder

**DOI:** 10.1101/2022.02.11.22270724

**Authors:** Viktoria-Eleni Gountouna, Mairead L. Bermingham, Ksenia Kuznetsova, Daniel Urda Muñoz, Felix Agakov, Siân E. Robson, Joeri J. Meijsen, Archie Campbell, Caroline Hayward, Eleanor M. Wigmore, Toni-Kim Clarke, Ana Maria Fernandez, Donald J. MacIntyre, Paul McKeigue, David J. Porteous, Kristin K. Nicodemus

## Abstract

Depression is a common psychiatric disorder with substantial recurrence risk. Accurate prediction from easily collected data would aid in diagnosis, treatment and prevention. We used machine learning in the Generation Scotland cohort to predict lifetime risk of depression and, among cases, recurrent depression. Rank aggregation was used to combine results across ten different algorithms and identify highly predictive variables. The model containing all but the cardiometabolic predictors had the highest predictive ability on independent data. Rank aggregation produced a reduced set of predictors without decreasing predictive performance (lifetime: 20 out of 154 predictors and Receiver Operating Characteristic area under the curve (AUC)=0·84, recurrent: 10 out of 180 predictors and AUC=0·76). Here we develop a pipeline which leads to a small set of highly predictive variables. This information can be easily collected with a smartphone ‘application’ to help diagnosis and treatment, while longitudinal tracking may help patients in self-management.

**Significance:** Depression is the most common psychiatric disorder and a leading cause of disability worldwide. Patients are often diagnosed and treated by non-specialist clinicians who have limited time available to assess them. We present a novel methodology which allowed us to identify a small set of highly predictive variables for a diagnosis of depression, or recurrent depression in patients. This information can easily be collected using a tablet or smartphone application in the clinic to aid diagnosis.

## Introduction

Major depressive disorder (MDD) is one of the most common mental disorders with a lifetime prevalence of around 15% (1), and it is also frequently recurrent (2-5) or persistent. The World Health Organization predicts that by 2030 13% of the total global disease burden will be accounted for by depression (6). Reducing the burden of MDD is a key public health challenge for the 21st century (7), with accurate diagnosis and prediction of recurrence of critical importance in reducing the disease burden. However, algorithms to predict incident and recurrent MDD that include data above that obtained on the basis of a structured clinical interview have contributed to improved prediction accuracies and may also improve diagnosis (8-10), especially when many patients see non-specialist clinicians such as general practitioners for diagnosis and treatment, where structured clinical interviews are impractical due to time constraints.

Prediction algorithms using standard statistical methodology have been developed previously for incident and/or recurrent MDD (7-12). These algorithms generated nominal to fairly high discriminative accuracy up to a C-statistic or AUC value of 0·79 (8-10). In these previous studies, the number and type of available predictors were limited to a small number of clinical features and applied standard statistical methodologies to develop prediction algorithms. However, in the age of “Big Data” – large-scale biobanking efforts with large sample sizes and hundreds to thousands of potential predictors – these standard statistical methodologies may be limited in their ability to advance personalized medicine in high-dimensional data. However, for clinical utility it is crucial to identify a concise and easily measured set of predictors with high predictive ability which can be used to determine a patient’s risk for lifetime or recurrent MDD. Ideally, these data could be combined with electronic health records to improve prediction and increase clinical utility. Indeed, advanced approaches such as machine learning have been successfully applied to prediction of treatment outcome in MDD (13).

We applied a set of modern “Big Data” approaches – a set of state-of-the-art machine learning methodologies – to the Generation Scotland: Scottish Family Health Study (GS:SFHS) cohort to predict lifetime and recurrent MDD. The predictors in GS:SFHS include cognitive function, personality, health and family history, socio-demographic, biometric, clinical and genomic data (14-15). We sought to apply these algorithms to a set of nested models to assess (a) algorithm performance and (b) to define a sparse set of predictors that may be useful in prediction in clinical practice by using a novel approach: the Markov Chain 4 (MC4) algorithm, which was developed for rank aggregation of Internet search engine rankings (16).

## Results

### Design Framework

We employed an array of machine learning algorithms, including tree-based, regression-based, neural networks and support vector machines (Methods). Data was divided into training and test sets (Fig. 1a). The training data was used to optimise hyperparameters for each algorithm using ten-fold cross validation and build prediction models (Fig. 1b). We assessed the performance of four nested models, described in detail in the Methods. Rank aggregation was used on the variable importance measures from the training data to select the top variables across methods. We performed independent replication on the test data using the models built on the training data with the area under the ROC curve as an outcome of interest.

**Fig. 1.**
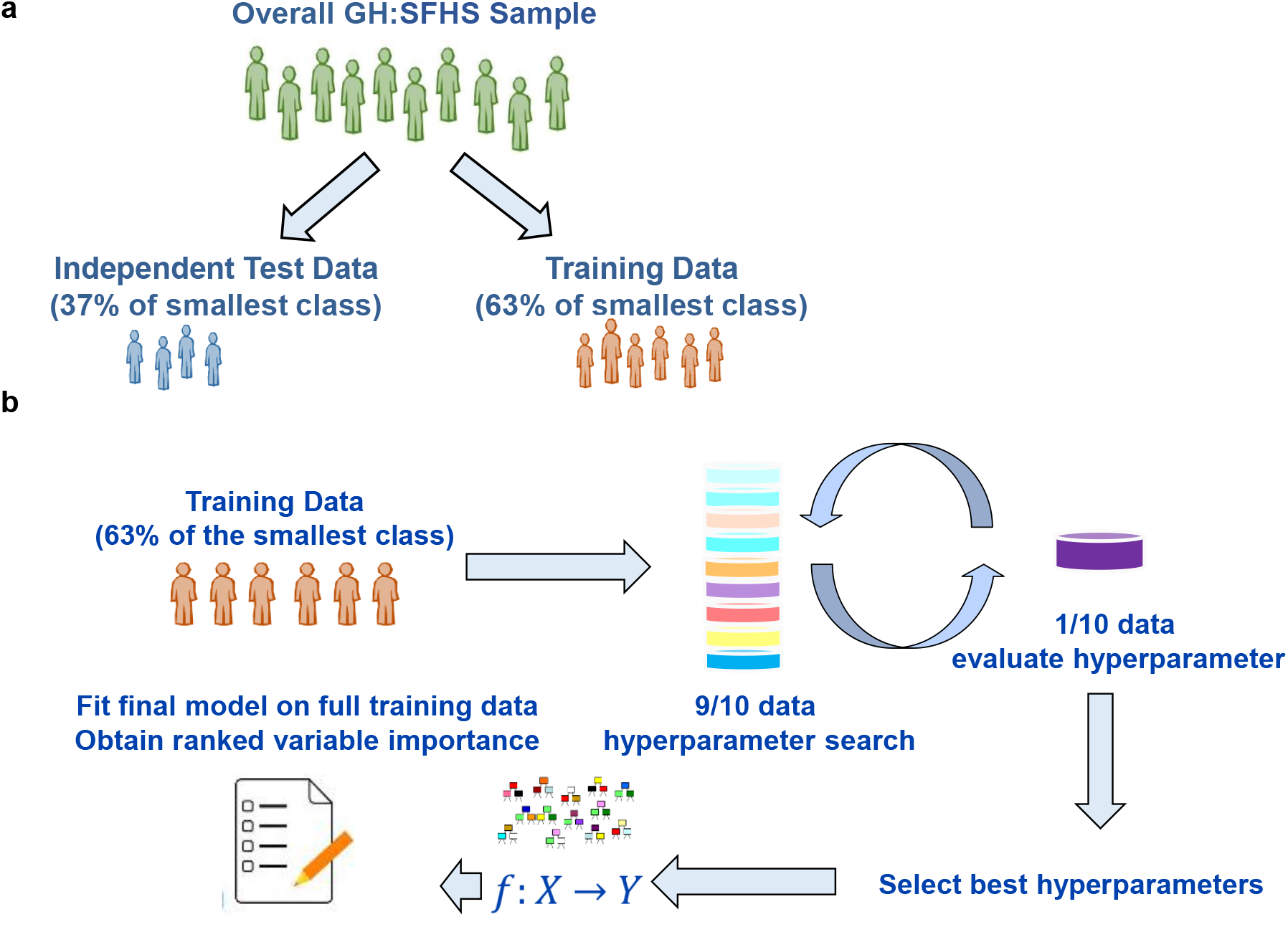
Schematic of Study Design. (a). Training and test sample creation. (b). Overall schematic of the study design. First, the training data are put through 10-fold cross validation to estimate appropriate hyperparameters. Second, the correct hyperparameters are applied to the full training set to create a model for prediction, and the variable importance measure rankings are recorded. Finally, the models developed on the training data are used for prediction of either lifetime MDD or single versus recurrent MDD on the test data.

### Demographics

For lifetime MDD, in both the training and test data, individuals with MDD were significantly younger, more likely to be female, to not be living as a couple, to live alone, to report lower income and live in areas that are more socioeconomically deprived than controls (all *p*-values < Bonferroni-corrected threshold of 0·005; SI Appendix, Table S1). There were no significant demographic differences between single and recurrent cases in the training or test sets using the Bonferroni *p*-value threshold.

### Predictive Performance

The predictive performance across the four models and machine learning algorithms for lifetime and recurrent MDD are summarised in Table 1 for lifetime MDD and Table 2 for single versus recurrent MDD. In both study designs, model one performed the poorest, model two increased predictive performance, and model three outperformed all other models. Model four, including cardiometabolic predictors, did not significantly improve upon model three; in fact, for some algorithms the inclusion of these additional variables led to significantly decreased predictive ability. For the best-performing algorithm in lifetime MDD, gradient descent boosting (GDB), model three increased the AUC versus model two from approximately 0·71 to 0·84, which is a more than threefold increase in information for discrimination, from 0.4 bits to 1.4 bits, although AUC values were slightly lower than what is commonly used for biomarkers (17). This improvement was also observed for recurrent MDD, although AUC values for Model two were smaller, less than 0·6. The increase in AUC for Model three for the best model, random forest (RF), increased the AUC from Model two of 0·58 to around 0·76.

**Table 1.** AUC Results for Predicting MDD on Independent Test Data: Lifetime MDD.

| Algorithm | M1 AUC | M1 95% CI | M2 AUC | M2 95% CI | M3 AUC | M3 95% CI | M4 AUC | M4 95% CI |
| --- | --- | --- | --- | --- | --- | --- | --- | --- |
| C5.0 | 0.652 | (0.625, 0.679) | 0.694* | (0.666, 0.723) | 0.813** | (0.789, 0.837) | 0.805 | (0.780, 0.830) |
| Conditional inference forest | 0.670 | (0.642, 0.697) | 0.709* | (0.681, 0.737) | 0.825**** | (0.802, 0.848) | 0.829* | (0.806, 0.852) |
| Elastic net | 0.670 | (0.643, 0.697) | 0.705* | (0.677, 0.734) | 0.835**** | (0.812, 0.858) | 0.834 | (0.811, 0.857) |
| Forward stepwise regression | 0.669 | (0.642, 0.697) | 0.711* | (0.683, 0.739) | 0.829**** | (0.805, 0.853) | 0.820 | (0.795, 0.844) |
| Gradient descent boosting | 0.663 | (0.635, 0.69) | 0.712** | (0.683, 0.741) | **0.839\*\*\*** | **(0.816, 0.861)** | 0.840 | (0.819, 0.862) |
| Neural networks | 0.666 | (0.640, 0.694) | 0.709* | (0.681, 0.736) | 0.824**** | (0.801, 0.847) | 0.825 | (0.801, 0.849) |
| Random forest | 0.660 | (0.632, 0.688) | 0.710* | (0.681, 0.739) | 0.824**** | (0.801, 0.848) | 0.828 | (0.804, 0.851) |
| SVM, linear | 0.666 | (0.639, 0.694) | 0.704* | (0.675, 0.734) | 0.833**** | (0.809, 0.856) | 0.830 | (0.807, 0.854) |
| SVM, polynomial | 0.670 | (0.639, 0.694) | 0.704* | (0.675, 0.733) | 0.830**** | (0.809, 0.856) | 0.831 | (0.807, 0.854) |
| SVM, radial basis function | 0.652 | (0.623, 0.680) | 0.694* | (0.665, 0.722) | 0.829**** | (0.806, 0.853) | 0.830 | (0.806, 0.853) |

**Table 2.** AUC Results for Predicting MDD on Independent Test Data: Single versus Recurrent MDD.

| Algorithm | M1<br>AUC | M1 95%<br>CI | M2<br>AUC | M2 95% CI | M3 AUC | M3 95%<br>CI | M4<br>AUC | M4 95%<br>CI |
| --- | --- | --- | --- | --- | --- | --- | --- | --- |
| C5.0 | 0.555 | (0.518,<br>0.592) | 0.536 | (0.487,<br>0.585) | 0.689** | (0.644,<br>0.734) | 0.650* | (0.584,<br>0.681) |
| Conditional inference forest | 0.593 | (0.543,<br>0.644) | 0.603 | (0.554,<br>0.653) | 0.728** | (0.686,<br>0.770) | 0.742* | (0.701,<br>0.783) |
| Elastic net | 0.584 | (0.532,<br>0.636) | 0.601 | (0.552,<br>0.650) | 0.742** | (0.700,<br>0.784) | 0.739 | (0.696,<br>0.782) |
| Forward stepwise regression | 0.589 | (0.537,<br>0.640) | 0.570 | (0.520,<br>0.619) | 0.699* | (0.655,<br>0.742) | 0.651* | (0.604,<br>0.698) |
| Gradient descent boosting | 0.592 | (0.541,<br>0.643) | 0.600 | (0.549,<br>0.648) | 0.756*** | (0.715,<br>0.796) | 0.735* | (0.692,<br>0.778) |
| Neural networks | 0.587 | (0.538,<br>0.636) | 0.557 | (0.507,<br>0.606) | 0.749*** | (0.706,<br>0.792) | 0.713* | (0.668,<br>0.757) |
| Random forest | 0.544 | (0.495,<br>0.594) | 0.576 | (0.526,<br>0.626) | **0.759\*\*\*** | **(0.718,**<br>**0.800)** | 0.743 | (0.700,<br>0.787) |
| SVM, linear | 0.587 | (0.535,<br>0.639) | 0.592 | (0.543,<br>0.641) | 0.695* | (0.650,<br>0.740) | 0.667* | (0.619,<br>0.715) |
| SVM, polynomial | 0.584 | (0.531,<br>0.636) | 0.595 | (0.546,<br>0.644) | 0.709* | (0.665,<br>0.754) | 0.697 | (0.652,<br>0.742) |
| SVM, radial basis function | 0.557 | (0.507,<br>0.606) | 0.571 | (0.522,<br>0.621) | 0.705** | (0.660,<br>0.750) | 0.692 | (0.646,<br>0.738) |

M = Model, AUC = Receiving Operator Characteristic Area Under the Curve, CI = Confidence Interval. The best performing model is denoted in bold type. \* $p < 0.05$ , \*\* $p < 1 \times 10^{-05}$ , \*\*\* $p < 1 \times 10^{-10}$ , \*\*\*\* $p < 1 \times 10^{-15}$ versus previous model.

After adjusting for 45 tests within each model and focusing on Model three as the best performing model, the GDB AUC was significantly higher than both C5.0 and conditional inference forest (CIF; *p*-values 0·00017 and 0·000028, respectively; SI Appendix, Table S2); other algorithms performed as well as GDB. For single versus recurrent MDD, C5.0 and linear support vector machine (SVM-L) showed significantly poorer performance versus the best-performing algorithms (SI Appendix, Table S3).

### Ranked Variable Importance and MC4 Analysis

The MC4 aggregate-ranked variable importance measures were used to calculate AUC for both lifetime and recurrent MDD, selecting ten, 20, 30…N of the ranked list to determine the minimum number of variables required to attain maximum AUC values for prediction of MDD in the independent test set. For lifetime MDD, this value was reached after the addition of the top 20 variables. The top ranked variables included (in order): neuroticism, General Health Questionnaire (GHQ) total score, GHQ depression, GHQ somatic symptoms, age, family history of depression, income, live as a couple, sex, mother with depression, whether the participant owned their home, GHQ anxiety, ever smoked, Mood Disorder Questionnaire (MDQ) score, Schizotypal Personality Questionnaire (SPQ) total score, educational qualification, pain intensity, GHQ social dysfunction, whether they ever had chronic pain, and if they live with someone else or live alone (Fig. 2a). For recurrent MDD, the maximum AUC was achieved after including only ten of the top-ranked variables. The top-ranking variables included (in order of rank): age at MDD onset, neuroticism, GHQ total score, digit symbol substitution (processing speed), GHQ somatic symptoms, GHQ depression, age, GHQ social dysfunction, whether they own their home and age when starting smoking (Fig. 2b). We observed considerable overlap between the two sets of predictors, including GHQ total and subscale depression, somatic symptoms and depression scores, neuroticism, age, and whether the participant owns their home. However, there were interesting differences, where for lifetime MDD family history of depression, MDQ and SPQ scores were critical predictors that were not included in the models predicting recurrent depression; and for recurrent depression processing speed was ranked highly by MC4, which was not included in the set of predictors for lifetime depression.

**Fig. 2.**
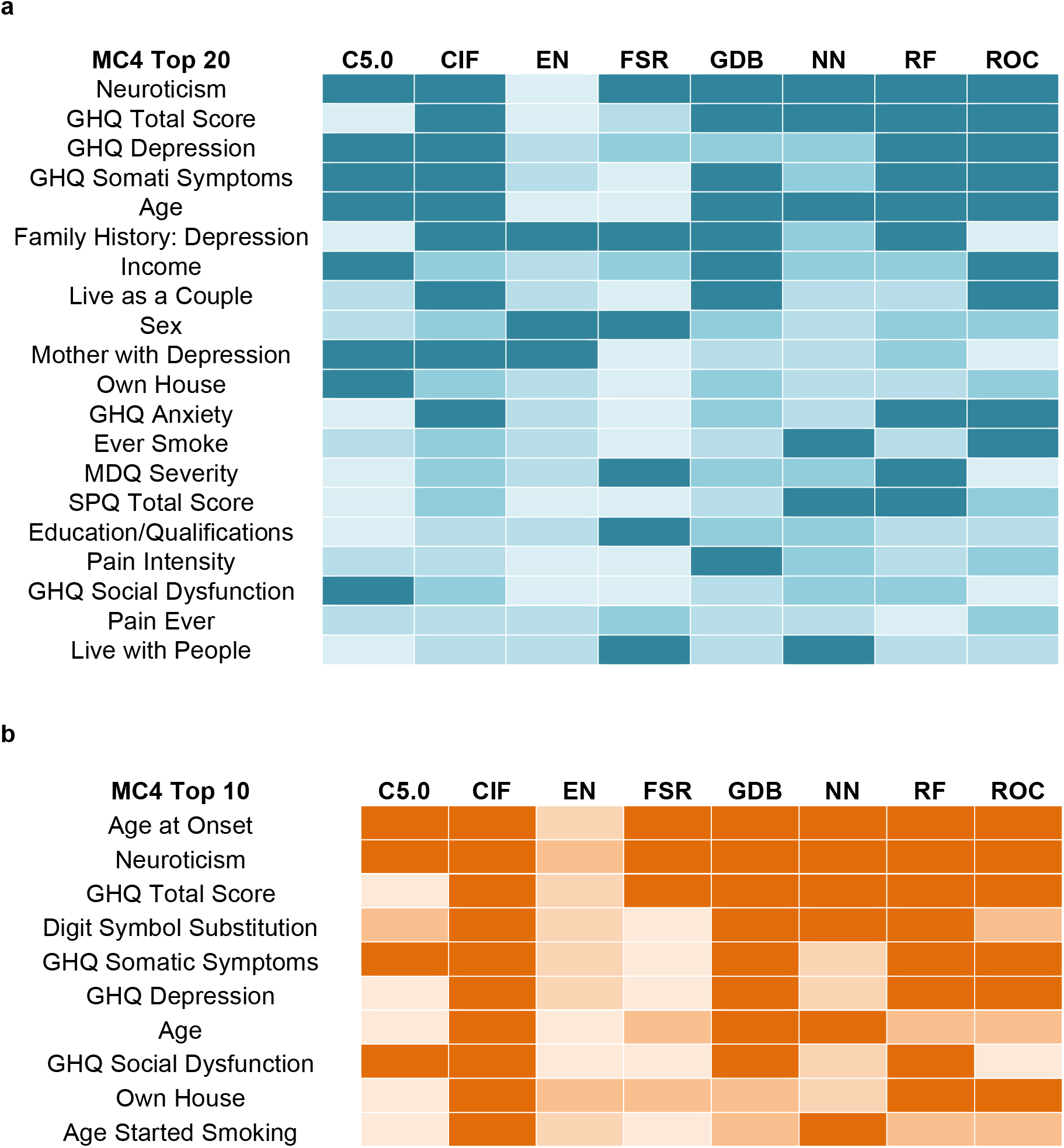
Concordance of individual algorithm rankings versus MC4 overall ranking. (a) Top panel shows concordance in lifetime MDD. Darkest blue = individual methods ranked in top 10, next darkest blue = top 20, light blue = top half, lightest blue = bottom half. The number of predictors in C5.0 (52) and FSR (41) was less than the top half of predictors, so any predictor with a zero-valued VIM was set to dark blue. GHQ = General Health Questionnaire, MDQ = Mood Disorder Questionnaire, SPQ = Schizotypal Personality Questionnaire (b) Bottom panel shows concordance in single versus recurrent episode MDD. Darkest orange = individual methods ranked in top 10, next darkest orange = top 20, light orange = top half, lightest orange = bottom half. The number of predictors in C5.0 (40), EN (37) and FSR (30) was less than the top half of predictors, so any predictor with a zero-valued VIM was set to dark orange. GHQ = General Health Questionnaire.

A comparison of the MC4-ranked variables and variables ranked by individual algorithms (Fig. 2a and 2b) showed that, with the exception of CIF in recurrent depression, none of the methods were fully consistent with the aggregated ranking. In fact, EN only ranked three predictors in the top 20 for lifetime depression and none in the top ten for recurrent depression. However, using the MC4-ranked subset of 20 variables for lifetime MDD and ten variables for single episode versus recurrent MDD led to equal – if not improved – performance across all methods. In lifetime MDD, the MC4-based model AUC values did not differ from those using the full set of 154 variables (Table 2). In single versus recurrent MDD, the reduced set of ten variables produced from MC4 improved predictive performance at the *p*-value < 0·05 level across six of the ten methods, for example, increasing the AUC for SVM-L from 0·695 using 180 variables in Model three to 0·771 using the MC4 set of ten.

### Leave-One-Out Analysis of MC4 Sets

To assess the relative contribution of predictors in the MC4 sets, we performed leave-one-variable-out analysis of these subsets, holding the random number seed constant. For lifetime MDD, only the removal of neuroticism showed a significant reduction in AUC values across machines (Δ AUC range = 0·024-0·034; *p*-values range = 0·00018 – 2·80 × 10^−08^; Supplementary Table 4). For single versus recurrent MDD, the removal of age at onset showed the largest reduction in AUC values across algorithms, with a change in AUC ranging from 0.052 for CIF to 0.088 for C5.0 (significant *p*-value range = 0·00052 to 2·36×10^−05^). Other variables showed a variable pattern across methods when being removed; none showed a consistent increase or decrease, which was expected as MC4 aggregates rankings across methods. However, the change in AUC values were small for all other variables.

### Effect size of Model 3 and MC4 Models

Hedges *g* was used to estimate effect sizes across different algorithms for Model three and the MC4 model in both study designs (Supplementary Table 6). Lifetime effect sizes were large, with all *g* values > 1·0 (Model three range = 1·06 to 1·41 MC4 range = 1·05 to 1·26). For single versus recurrent MDD, the use of the MC4 set increased *g* values; in fact, for several algorithms the *g* values were within the range of “medium” (e.g., < 0·8) when using Model three, but all *g* values were large (ranging from 0·93 to 1·13) using MC4 (Supplementary Table 6).

## Discussion

Using state-of-the-art machine learning methods combined with a novel usage of the MC4 algorithm, we have defined a set of predictive variables with AUC values that are larger than those observed in previous studies; for lifetime MDD, the AUC value is within the range of predictive ability for use in clinic. These algorithms were trained on a large biobank with deep phenotyping, in contrast to previous approaches, and may be easily applied to other similar “Big Data” applications.

The most consistent demographic factors associated with lifetime MDD risk in previous studies have been female sex, younger age and low socioeconomic status (18-22). Psychosocial risk factors commonly associated with MDD include negative life events, traumatic experiences, work-related stress, financial strain, poor marital or interpersonal relationships, lack of social support and low self-esteem (10, 23-26). The number of previous episodes, the level of residual symptoms, and childhood maltreatment are consistent risk factors of recurrent MDD (27-28). We highlight that our findings suggest neuroticism is one of the top predictors of lifetime MDD and also recurrent MDD. Neuroticism is easily measured by a general practitioner in clinic and would be the best single measure for prediction of both types of depression.

Using our algorithm, we provide descriptive characteristics of individuals who are at each quintile of our prediction engine, as an illustration of patients a general practitioner may see in clinic (Table 3). For example, versus individuals who would have the lowest predicted risk of lifetime MDD (Table 4), individuals at higher risk levels would be more likely to score higher on measures of neuroticism, general psychological distress (GHQ), have a family history of depression, be a current smoker and live alone. For individuals with a higher predicted probability of having recurrent depression versus single-episode (Table 5), higher risk for recurrent MDD would be more likely observed in individuals with a lower age at MDD onset, higher neuroticism, higher psychological distress (GHQ), and poorer performance on digit symbol coding.

**Table 3.** AUC Results for Best Performing Model versus MC4 Model.

| Algorithm | Lifetime MDD: M3 = 154 Variables,<br>MC4 = 20 Variables |  |  |  | Recurrent MDD: M3 = 180<br>Variables, MC4 = 10 Variables |  |  |  |
| --- | --- | --- | --- | --- | --- | --- | --- | --- |
|  | M3<br>AUC | M3 95%<br>CI | MC4<br>AUC | MC4 95%<br>CI | M3<br>AUC | M3 95%<br>CI | MC4<br>AUC | MC4<br>95% CI |
| C5.0 | 0.813 | (0.789,<br>0.837) | 0.808 | (0.784,<br>0.833) | 0.689 | (0.644,<br>0.734) | 0.735* | (0.694,<br>0.776) |
| Conditional inference forest | 0.825 | (0.802,<br>0.848) | 0.825 | (0.802,<br>0.848) | 0.728 | (0.686,<br>0.770) | 0.743* | (0.702,<br>0.784) |
| Elastic net | 0.835 | (0.812,<br>0.858) | 0.831 | (0.809,<br>0.854) | 0.742 | (0.700,<br>0.784) | 0.771* | (0.733,<br>0.809) |
| Forward stepwise regression | 0.829 | (0.805,<br>0.853) | 0.834 | (0.812,<br>0.857) | 0.699 | (0.655,<br>0.742) | 0.771* | (0.733,<br>0.809) |
| Gradient descent boosting | **0.839** | **(0.816,**<br>**0.861)** | **0.839** | **(0.817,**<br>**0.862)** | 0.756 | (0.715,<br>0.796) | **0.775\*** | **(0.736,**<br>**0.813)** |
| Neural networks | 0.824 | (0.801,<br>0.847) | 0.835 | (0.812,<br>0.858) | 0.749 | (0.706,<br>0.792) | 0.752 | (0.712,<br>0.792) |
| Random forest | 0.824 | (0.801,<br>0.848) | 0.830 | (0.808,<br>0.853) | **0.759** | **(0.718,**<br>**0.800)** | 0.762 | (0.721,<br>0.803) |
| SVM, linear | 0.833 | (0.809,<br>0.856) | 0.832 | (0.809,<br>0.855) | 0.695 | (0.650,<br>0.740) | 0.771* | (0.732,<br>0.809) |
| SVM, polynomial | 0.830 | (0.809,<br>0.856) | 0.832 | (0.809,<br>0.854) | 0.709 | (0.665,<br>0.754) | 0.740 | (0.698,<br>0.782) |
| SVM, radial basis function | 0.829 | (0.806,<br>0.853) | 0.827 | (0.805,<br>0.850) | 0.705 | (0.660,<br>0.750) | 0.743 | (0.700,<br>0.786) |

M = Model, AUC = Receiving Operator Characteristic Area Under the Curve, CI = Confidence Interval. The best performing models are denoted in bold type. \* $p < 0.05$ , \*\* $p < 1 \times 10^{-05}$ , \*\*\* $p < 1 \times 10^{-10}$ , \*\*\*\* $p < 1 \times 10^{-15}$ for comparison of model 3 to MC4 model.

**Table 4.** Examples of Individuals at Quintiles of Predicted MDD using MC4 Predictors: Lifetime MDD.

| Prediction Quintile | Neu | GHQ Total | GHQ-Dep | GHQ-Som | Age | Fx Dep | Income Level | Live as Couple | Sex | Mother Depress | Own House | GHQ-Anx | Ever Smoke | MDQ-C | SPQ-Total | Ed/Qual | Pain Inten | GHQ-Soc | Pain Ever | Number in House |
| --- | --- | --- | --- | --- | --- | --- | --- | --- | --- | --- | --- | --- | --- | --- | --- | --- | --- | --- | --- | --- |
| 0% | 0 | 7 | 0 | 0 | 65 | No | Lowest | Yes | M | No | Yes | 0 | No | None | 1 | School Leavers Certificate | 40 | 7 | No | 2 |
| 25% | 0 | 5 | 0 | 1 | 61 | No | Highest | No | M | No | Yes | 0 | Current | None | 5 | University | 38 | 4 | Yes | 1 |
| 50% | 4 | 14 | 0 | 5 | 64 | No | Second-Highest | Yes | F | No | Yes | 2 | No | None | 7 | University | 21 | 7 | Yes | 2 |
| 75% | 4 | 11 | 0 | 2 | 49 | No | Highest | No | M | No | Yes | 6 | No | Serious | 3 | University | 56 | 3 | Yes | 3 |
| 100% | 8 | 22 | 4 | 5 | 25 | Yes | Highest | No | F | No | No | 10 | Current | Serious | 12 | University | 33 | 3 | Yes | 1 |

**Table 5.**
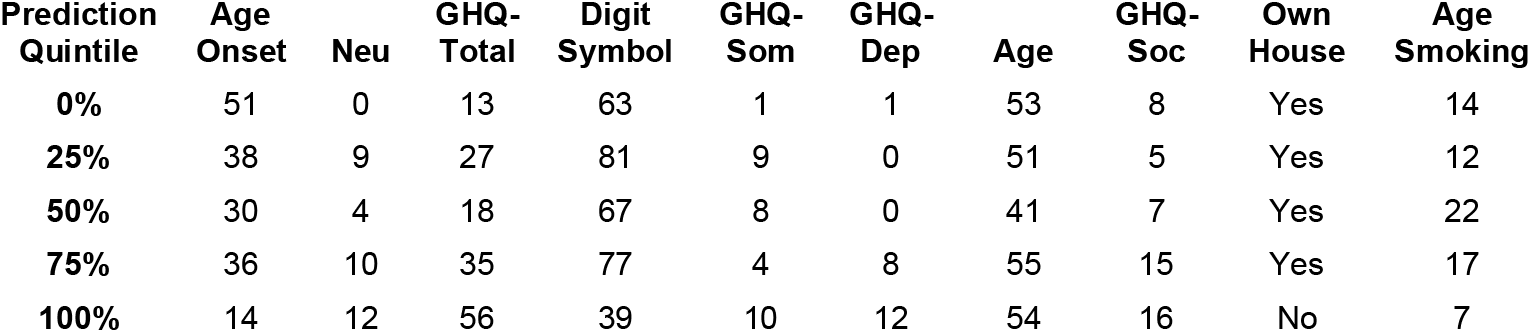
Examples of Individuals at Quintiles of Predicted MDD using MC4 Predictors :Single versus Recurrent MDD.

| Prediction Quintile | Age Onset | Neu | GHQ-Total | Digit Symbol | GHQ-Som | GHQ-Dep | Age | GHQ-Soc | Own House | Age Smoking |
| --- | --- | --- | --- | --- | --- | --- | --- | --- | --- | --- |
| 0% | 51 | 0 | 13 | 63 | 1 | 1 | 53 | 8 | Yes | 14 |
| 25% | 38 | 9 | 27 | 81 | 9 | 0 | 51 | 5 | Yes | 12 |
| 50% | 30 | 4 | 18 | 67 | 8 | 0 | 41 | 7 | Yes | 22 |
| 75% | 36 | 10 | 35 | 77 | 4 | 8 | 55 | 15 | Yes | 17 |
| 100% | 14 | 12 | 56 | 39 | 10 | 12 | 54 | 16 | No | 7 |

The main strength of this study is that it has assessed the importance and contribution of well-known and novel predictors of MDD measured using standardised tests in a large population-based cohort; and further introduces the MC4 algorithm for rank aggregation of results. Generation Scotland is a large homogeneous population that to date is one of the few cohorts containing a clinical definition of MDD on a large-scale sample. The two study designs (lifetime MDD–healthy controls and single episode-recurrent MDD) allow for insights into shared and differing risk factors. Limitations of this study are the sample size and the lack of replication in an independent large-scale cohort with a clinical definition of MDD.

This study has shown that a combination of simple self-report measures can form the basis of models that provide excellent prediction of lifetime or recurrent MDD. Given that the measures identified through the MC4 algorithm are relatively easy to obtain, a possible implementation of the findings of this work could be to collect these measures at regular intervals, which would provide a basis for longitudinal assessment of change in individuals or groups of patients. Such evidence on patient outcomes would be useful in tracking the impact of interventions, especially treatment response to antidepressants or psychological interventions such as cognitive behavioural therapy. If incorporated into clinical practice, the use of algorithms to predict a patient’s risk of MDD could have a substantial impact on the timeliness and effectiveness of diagnosis and treatment. The measures can be obtained outside of the clinic and the resulting data-based risk prediction can inform clinicians’ decisions, allowing more time during consultations to discuss issues specific to the patient to accommodate a personalised approach. Another exciting prospect for the implementation of the results of this study could therefore be to use the MC4 algorithm as the basis of a decision support tool that could be incorporated into a mobile application (app) for the benefit of patients, support workers and clinicians. Patients could enter their own data into the app and a built-in algorithm would be able to forewarn or reassure about lifestyle changes that might presage or mitigate recurrence. Apps are increasingly being developed and used successfully in health and social care to access treatment guidelines and to support decisions about patient screening, treatment options and drug dosage (29). In the current era of increased availability of data, new systems for linking and utilising healthcare and other records could provide crucial and timely information to support personalised healthcare.

## Materials and Methods

### Generation Scotland: Scottish Family Health Study

Our sample was drawn from the GS:SFHS, a large population-based study. A full description of the cohort and protocol for recruitment is described in detail elsewhere (14-15). Briefly, the GS:SFHS is a representative survey of the Scottish population, consisting of 23,960 individuals over 18 years of age recruited between the years 2006 and 2011. All components of GS:SFHS received ethical approval from the NHS Tayside Committee on Medical Research Ethics (REC Reference Number: 05/S1401/89) and all participants gave written informed consent. Generation Scotland data are available under managed access by submitting a proposal to the Generation Scotland Data Access Committee.

The selection criteria for participants included: Caucasian ethnicity, born in the UK and phenotype data available from attendance at a Generation Scotland research clinic (30). We further restricted the analysis to unrelated individuals, as the machine learning algorithms employed required independent observations. Cases were not matched to controls as doing so would remove the effect of the matching criteria from analysis; thus preventing assessment of potential interaction effects for those criteria.

### Assessment of MDD

All participants in GS:SFHS were screened for psychiatric disorders and those who responded positively to screening questions were invited to continue with the MDD and bipolar disorder modules from the Structured Clinical Interview for Diagnostic and

Statistical Manual of Mental Disorders, Fourth Edition (SCID;31). The diagnostic interview was conducted in person by trained clinical research nurses.

### Predictors of MDD: Self-Report, Clinical, Cognitive and Demographic

#### MDD Symptomology

For analysis of single versus recurrent depression, we included 12 symptom predictors from the SCID interview, plus age at onset and a variable indicating which nurse conducted the interview. Missing values for SCID interview questions were controlled for by creating a binary vector indicating which values were not recorded.

#### General Psychological Distress

Psychological distress was measured using the 28-item General Health Questionnaire (GHQ; 32) at baseline by a trained clinical nurse. Fifty-seven percent of the study participants were also given the Mood Disorder Questionnaire (MDQ; 33) and Schizotypal Personality Questionnaire-Brief (SPQ-B; 34).

#### Cognitive Performance

Cognitive performance was assessed using measures of processing speed (Wechsler Digit Symbol Substitution Task; 35), verbal declarative memory (Wechsler Logical Memory Test; 36) both immediate and delayed, executive function measured with the letter-based phonemic verbal fluency test using the letters C, F, and L, each for one minute (37) and vocabulary (Mill Hill Vocabulary Scale; 38).

#### Demographic and Socioeconomic Status

The Scottish Index of Multiple Deprivation (SIMD; 39) 2009, matched to each participant’s postcode, was used to assess socioeconomic status. SIMD is a ranking based on seven domains: income, employment, health, education, geographic access, crime, and housing.

#### Clinical Measurements, Personal and Family Medical History

Clinical measures included height, weight, BMI, waist, hip, waist:hip ratio, percent body fat, systolic and diastolic blood-pressure (both mean of two measurements), respiratory function (forced expiratory volume in one second, forced vital capacity and forced expiratory flow; each measured three times) and blood sodium, potassium, urea, creatinine, glucose, total cholesterol and HDL-cholesterol levels (14). Participants were asked about whether they or their first-degree relatives had a history of depression, Parkinson disease, Alzheimer disease, diabetes, asthma, cardiovascular disease (including heart disease, hypertension and stroke), osteoarthritis, rheumatoid arthritis, hip fracture or common cancers (including breast, bowel, lung and prostate).

#### Alcohol and Tobacco Use

Participants were identified as current drinkers, former drinkers (either stopping greater than or less than 12 months) or never drinkers. Current alcohol consumption was characterised as either more or less than usual for each participant. Total consumption was measured in self-reported units of alcohol consumed in the previous week. Participants were classified into two groups: ever-smokers (current smokers and former smokers) and never smokers, with ever-smokers also asked at what age they started smoking and amount of tobacco consumption.

#### Chronic Pain

A validated self-report chronic pain questionnaire was used to assess pain severity based on its intensity and impact of pain on daily functioning in the previous three months^40^.

### Analytic Methods

#### Training and Test Datasets

One thousand and nineteen randomly-selected unrelated participants who had at least one episode of major depressive disorder were randomly selected as cases. Randomly-selected, healthy unrelated individuals (N = 3,994) with no lifetime diagnosis of psychiatric disorders served as controls. For analyses examining single versus recurrent major depressive disorder, we randomly resampled unrelated individuals from GS:SFHS to increase sample size (as controls were excluded), leading to a total of 1,198.

The final data sets were divided into (a) training data for machine learning hyper-parameter optimisation and model building and (b) test data (Fig. 1a). The test data were not used to develop any prediction models and formed an independent replication set. Many machine learning methods for binary classification require relatively equal numbers of cases and controls to assure optimised prediction (41). We performed a down-sampling procedure to balance the case-control ratio in the training data. The minority class (e.g., cases with MDD and recurrent MDD) were randomly split into a training set (63%) and an independent test set (37%). The majority class (e.g., controls and single episode MDD) was randomly under-sampled to make its frequency equal to the minority class for the training data (see Table 1 for Ns). Hedges g (42) was used to estimate effect sizes because of imbalance in the independent test sets.

#### Machine Learning Algorithms and Hyper-Parameter Settings

A panel of predictive machine learning algorithms was applied to the data. We applied the C5.0 tree-based algorithm (43-44), two tree-based ensemble algorithms (random forest, RF; 45-46) and conditional inference forest (CIF; 47-48), a gradient descent boosting algorithm (GDB; 49-50), elastic net, a regularised regression technique (EN; 51-52), feed-forward neural networks (NN; 53-54), and support vector machines (SVM; 55-57) with the following kernels: linear (SVM-L), polynomial (SVM-P) and radial basis function (SVM-R). We also used forward stepwise regression with Akaike’s Information Criterion (58). These methods were selected as best examples of a range of machine learning methodologies commonly applied to biologic data.

The R package caret (59) was used to perform ten-fold cross validation on the training data to set optimised values for the hyper-parameters for each algorithm (Fig. 1b; details about hyper-parameter selection in SI Appendix, Methods). All analyses were conducted in R version 3.2.4 (60).

#### Receiver Operating Characteristic (ROC) area under the curve (AUC) and Variable Importance Measures (VIMs)

The outcome of interest was the area under the ROC curve from the independent test data using models built on the training data. We used the nonparametric DeLong (61) method as implemented in pROC (62) to test for statistical differences between areas under the curve among independent test data. We sought to find the largest ROC AUC whilst minimising the number of variables required such that a smaller set could potentially be used for prediction by general practitioners. We also report an alternative measure to the AUC to aid in interpretation, where to quantify differences in predictive performance we use the expected weight of evidence as measured in bits (63). Machine learning algorithms generate measures, or VIMs, of the relative contribution of predictors in classifier construction, or a measure of the strength of association between predictor and outcome. For C5.0 the VIM used was the percent of observations in the nodes directly under the split for that variable; for RF and CIF it was the permutation-based VIM; for EN and FSR it was the absolute value of the coefficients, for NN and all SVM we used caret’s internal VIM function, which is not dependent on the algorithm and simply measures the AUC increase when including the variable over a null model as these methods do not provide a direct measure of variable importance.

#### Markov Chain 4 (MC4) Algorithm

We took ranked variable importance measures from the training data using the optimal set of hyper-parameters for each algorithm and applied the Markov Chain 4 (MC4) algorithm (16, 64) to provide an aggregated variable ranking. To begin, the MC4 algorithm considers a single set *U* that contains the union of the top *k* elements from all lists. For each pair *i* and *j* in the list (*i* ≠ *j*), set the Markov chain transition matrix *M* element *m*_*ij*_ to 1/|*U*| if ≥ 50% of the lists rank *j* above *i* and 0/|*U*| otherwise. If *i* and *j* are never contained in the same list, then *m*_*ij*_ = *m*_*ji*_ = 0.5/|*U*|, and let *m*_*ii*_ =. If ε is a small positive constant, it creates an ergodic transition matrix *M* by ((1-ε)\**m*) + ε/|*U*|. The resulting stationary probabilities are then used to create an aggregate ranking of the variables, with higher probabilities denoting a higher rank (i.e., the Markov process spent longer time in those states). The proposed novel use of meta-ranking technology provides robust inferences about the relative contribution to risk for MDD across different machines. We then selected the top ten, 20, …, N ranked variables and fitted them in the best-performing model to assess the number of variables required to reach maximum predictive ability. Leaving one predictor out at a time, we assessed the relative contribution of each predictor to the final model AUC values in the independent test set.

#### Models Assessed

We assessed performance of four nested models, where *outcome*_*k*_ was either (1) lifetime MDD versus controls or (2) single versus recurrent depression:

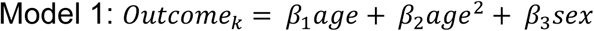

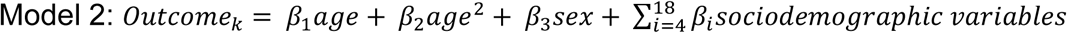

Building upon model two, the third model included mood, schizotypy and cognitive variables; family and personal medical history (excluding stroke, metabolic and cardiovascular history); as well as smoking and alcohol consumption. In addition, for the analysis of single versus recurrent MDD, we added variables derived from the SCID interview; this led to a total of 154 variables for lifetime MDD and 180 for analysis of single versus recurrent MDD. We hypothesised that cardiometabolic predictors may not improve prediction in all MDD cases but instead particularly in those with a later age at onset (65). Model four included Model three plus cardiovascular and stroke medical and family history; obesity, diabetes, FEV and lab measurements as these are commonly co-morbid with depression (N predictors: lifetime MDD: 211; single versus recurrent MDD: 237).

Hyperparameters were optimised using ten-fold cross validation on the training data. We used the nonparametric DeLong approach to test for statistical differences between areas under the curve among independent test data.

## Data Availability

Generation Scotland data are available under managed access by submitting a proposal to the Generation Scotland Data Access Committee.

https://www.ed.ac.uk/generation-scotland

## Author Contributions

KKN designed the study, supervised and performed analysis, and wrote the manuscript. VEG, MLB, and KK performed analysis and wrote the manuscript. JJM and SER wrote the manuscript. DJP co-designed the study and edited the manuscript. DUM, FA, PM, AC, CH, EW, TC, AMF, DJM performed analysis or assistance in data coding and edited the manuscript. AM edited the manuscript.

## Acknowledgments

VEG and KKN were supported by a University of Edinburgh-Medical Research Council (https://mrc.ukri.org/) Confidence in Concept award and by a Wellcome Trust (https://wellcome.ac.uk/)-University of Edinburgh Institutional Strategic Support Fund award (to KKN). KKN, MLB, DUM, FA, PM, DJP were supported by a University of Edinburgh-Medical Research Council (https://mrc.ukri.org/) Confidence in Concept award (to DJP). KKN and JJM were supported by the Rosetrees Trust (http://www.rosetreestrust.co.uk/; M405) and VEG, SER and KKN by a Chancellor’s Fellowship from the University of Edinburgh. DUM was supported by European Union FP7 (https://ec.europa.eu/research/fp7/index_en.cfm) 316861 MLPM. DJM acknowledges the financial support of NHS Research Scotland (http://www.nhsresearchscotland.org.uk/; NRS), through NHS Lothian. Generation Scotland received core support from the Chief Scientist Office of the Scottish Government Health Directorates (http://www.cso.scot.nhs.uk/) [CZD/16/6] and the Scottish Funding Council (http://www.sfc.ac.uk/) [HR03006]. Genotyping of the GS:SFHS samples was carried out by the Genetics Core Laboratory at the Wellcome Trust Clinical Research Facility, Edinburgh, Scotland and was funded by the Medical Research Council UK (https://mrc.ukri.org/) and the Wellcome Trust (https://wellcome.ac.uk/; Wellcome Trust Strategic Award “STratifying Resilience and Depression Longitudinally” (STRADL) Reference 104036/Z/14/Z). We are grateful to all the families who took part, the general practitioners and the Scottish School of Primary Care for their help in recruiting them, and the whole Generation Scotland team. The funders had no role in study design, data collection and analysis, decision to publish, or preparation of the manuscript.

## Supporting Information

### Supplementary Methods

#### Machine learning optimisation

The optimisation was set to a grid search for C5.0 (number of trials 1-20), CIF and RF (number of trees set to 1000, number of variables selected at each split (*mtry*) set to a grid of 10-15 values up to the total number of variables), GDB (interaction depth grid set to 1-3, number of iterations grid set to between 1 000 and 5 000, shrinkage held constant at 0.001), EN (grid for alpha and lambda set between 0 and 1). For NN and SVM models, we used a random search as the number of hyper-parameters was large and a random search has been shown to have equal or improved performance versus a grid search for these methods (1). Data were centred and scaled before analysis using SVMs as different scales of measurement can lead to numerical difficulties during the calculation of the inner products of the variables.

#### Imputation

For variables with less than 5% missing data, multivariate imputation by chained equations (MICE; 2-3) was used to replace missing values.

---

**Table S1.**
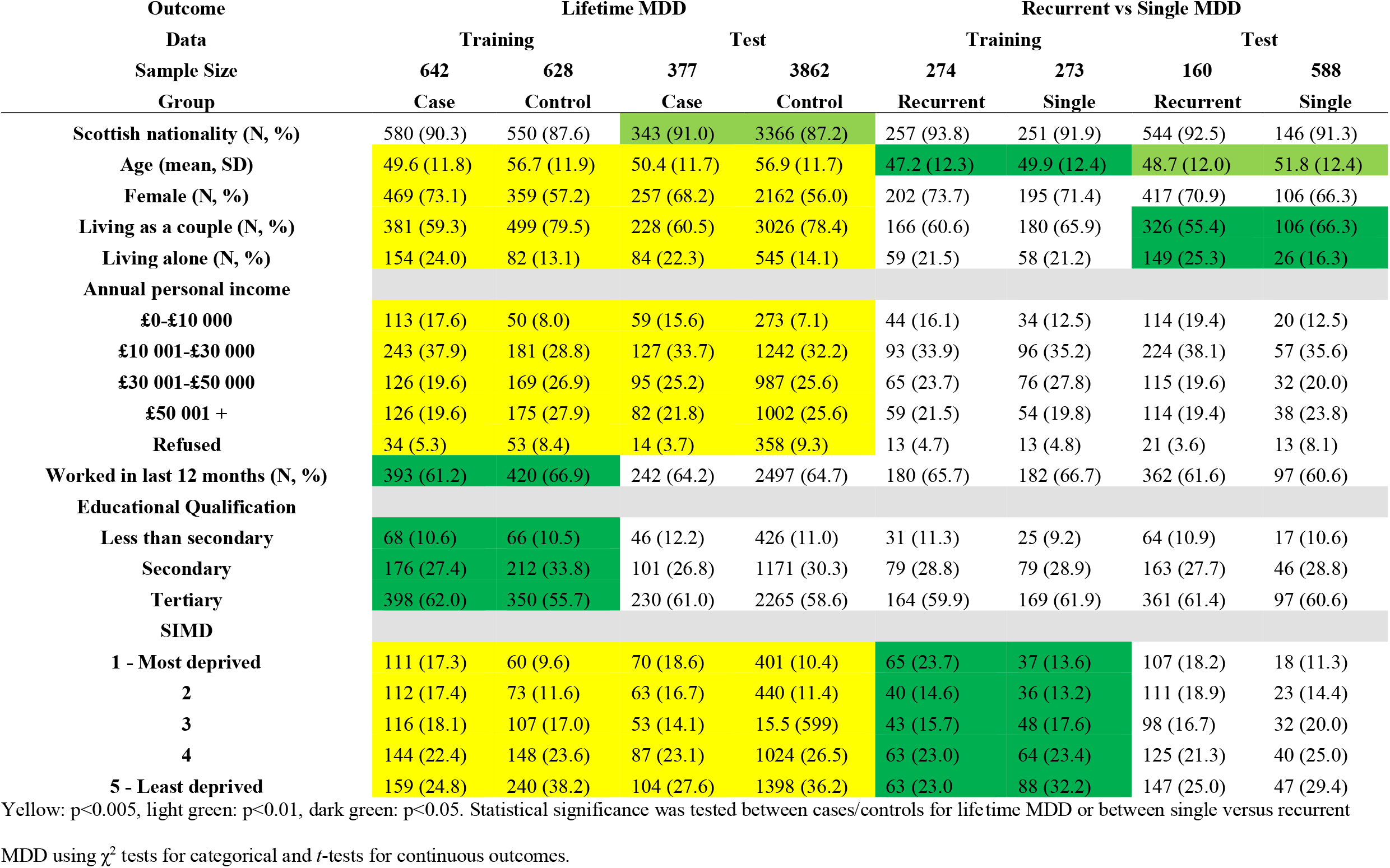
Demographic and Socioeconomic Characteristics in Cases and Controls and Single vs. Recurrent MDD.

**Table S2.**
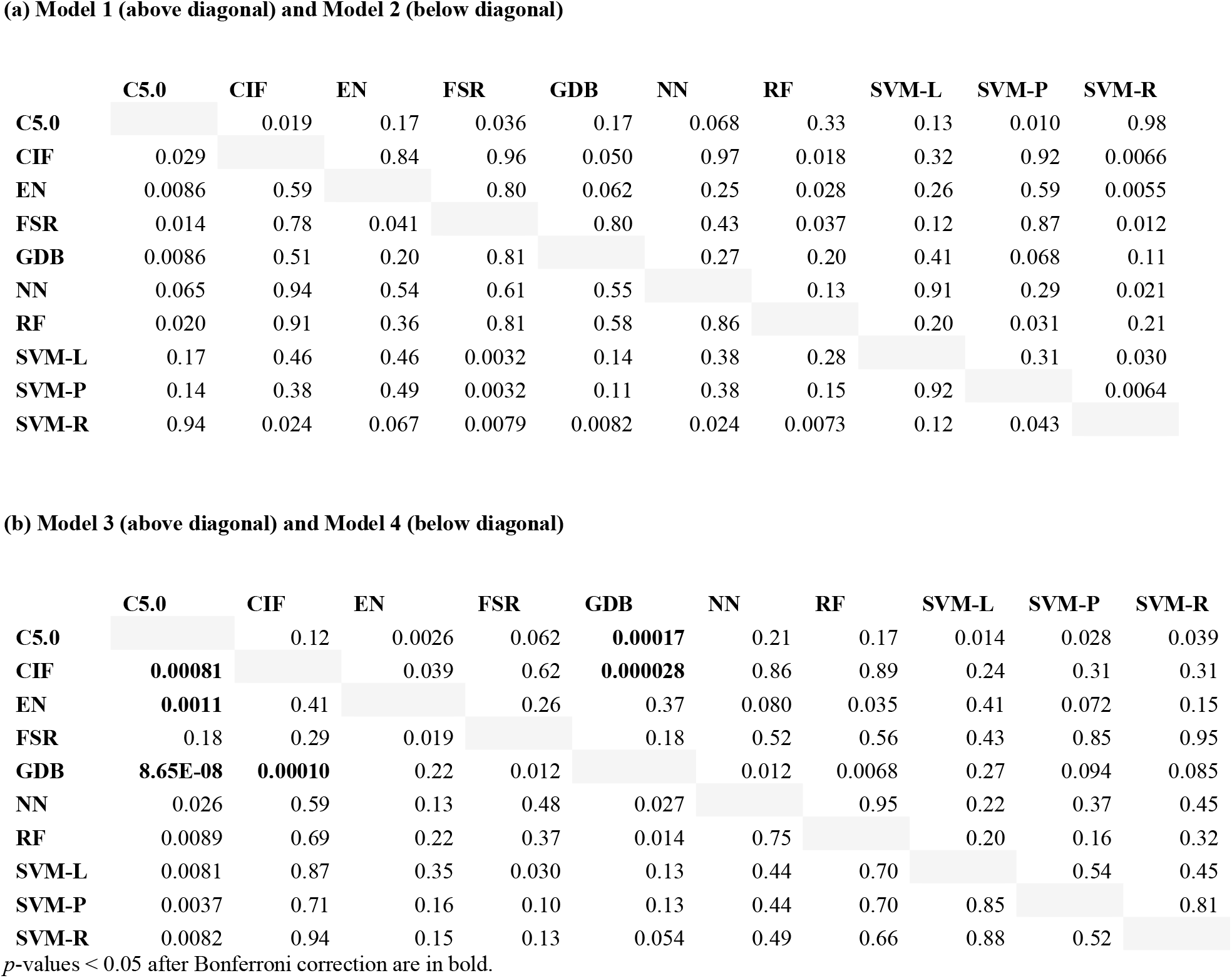
ROC Curve Comparisons between Methods within Models for Lifetime MDD.

**Table S3.**
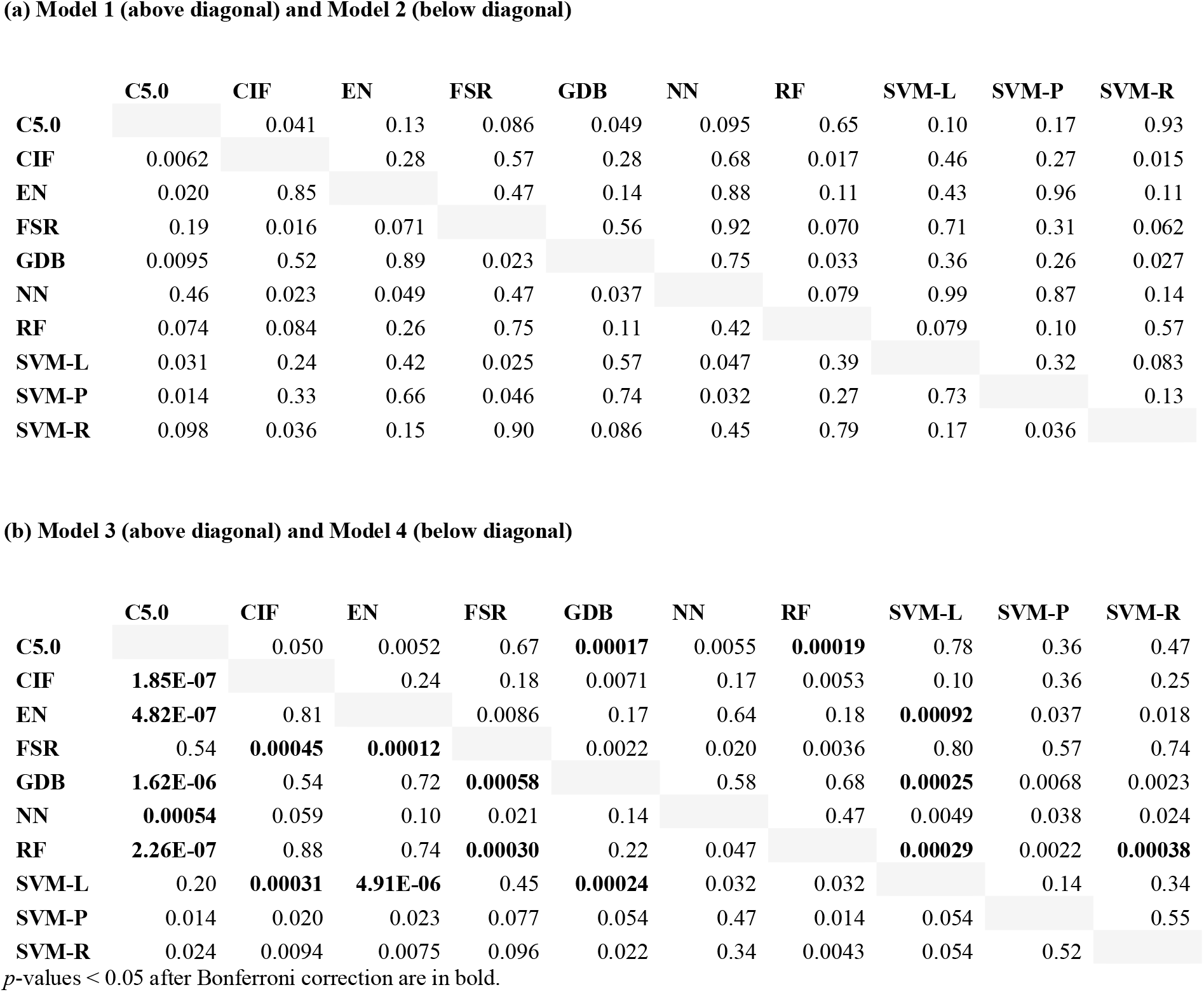
ROC Curve Comparisons between Methods within Models for Single versus Recurrent MDD.

**Table S4.**
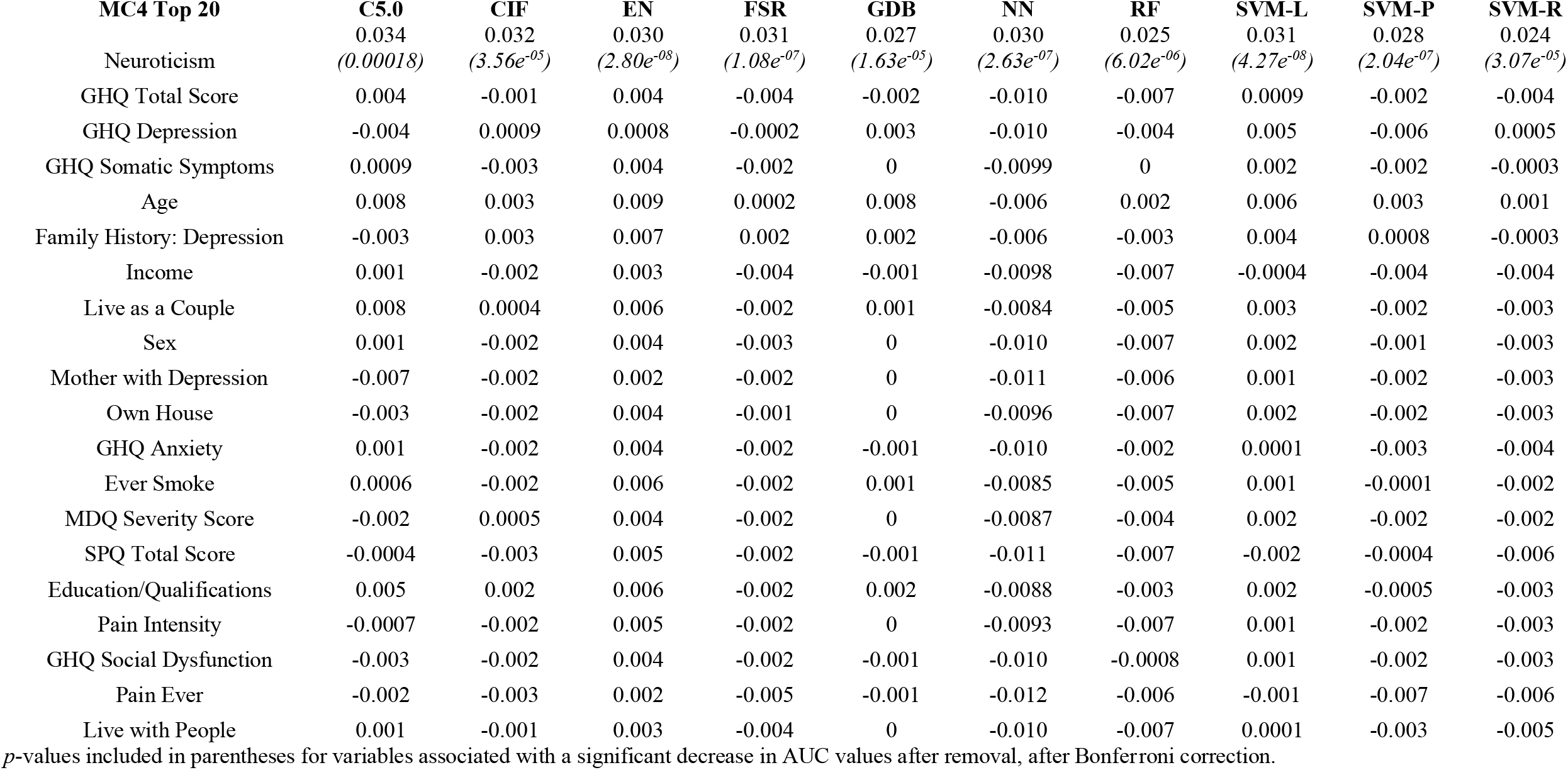
AUC Differences between MC4 Full Model and Leave-One-Out Analysis of Top 20 Predictors in Lifetime Depression.

**Table S5.**
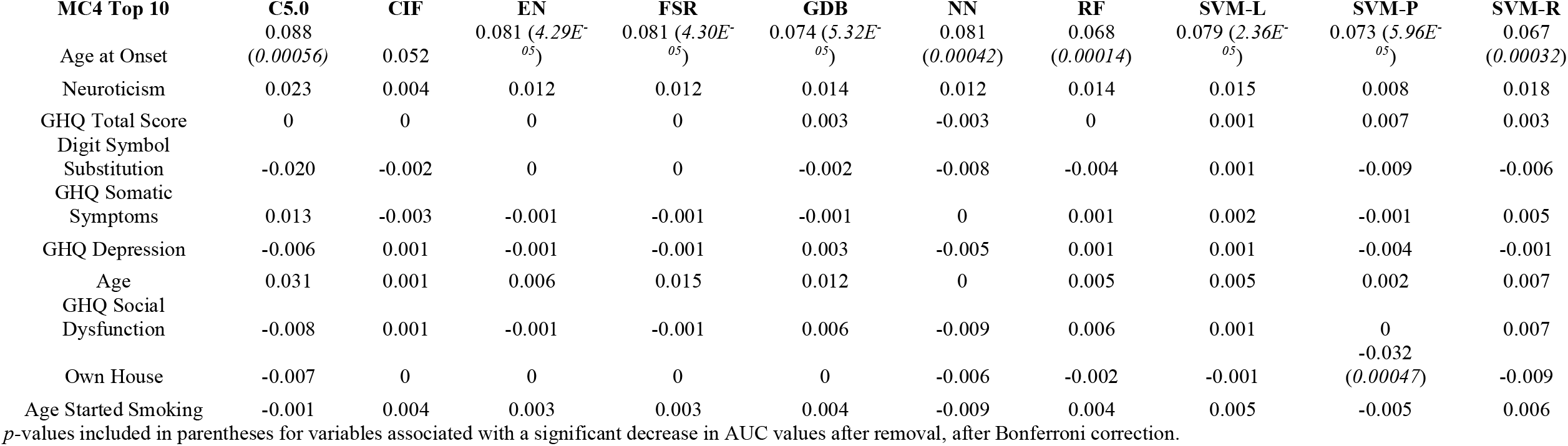
AUC Differences between MC4 Full Model and Leave-One-Out Analysis of Top 20 Predictors in Single versus Recurrent Depression.

**Table S6.**
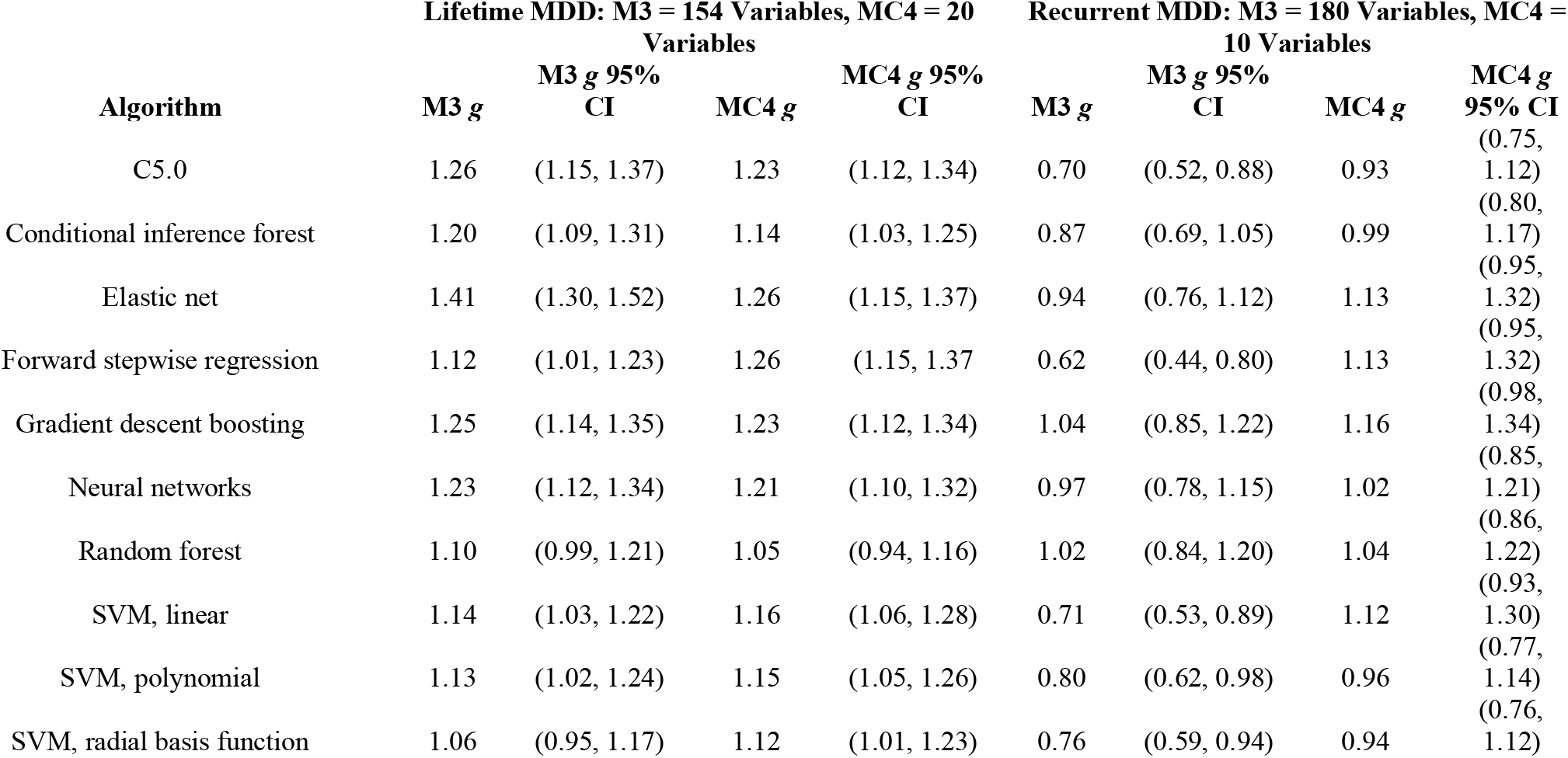
Effect Size Given by Hedges *g* for Model 3 and MC4 Model in Lifetime and Single versus Recurrent MDD.

