## Supplemental Materials for "Predictive Machine Learning for Personalised Medicine in Major Depressive Disorder"

**Supporting Information**

**Methods**

***Machine learning optimisation***

The optimisation was set to a grid search for C5.0 (number of trials 1-20), CIF and RF (number of trees set to 1000, number of variables selected at each split (*mtry*) set to a grid of 10-15 values up to the total number of variables), GDB (interaction depth grid set to 1-3, number of iterations grid set to between 1 000 and 5 000, shrinkage held constant at 0.001), EN (grid for alpha and lambda set between 0 and 1). For NN and SVM models, we used a random search as the number of hyper-parameters was large and a random search has been shown to have equal or improved performance versus a grid search for these methods (1). Data were centred and scaled before analysis using SVMs as different scales of measurement can lead to numerical difficulties during the calculation of the inner products of the variables.

***Imputation***

For variables with less than 5% missing data, multivariate imputation by chained equations (MICE; 2-3) was used to replace missing values.

**Supporting Information References**

1. Bergstra J, Bengio Y. Random Search for Hyper-Parameter Optimization. J. Mach. Learn. Res. 2012;13(Feb):281–305.

2. van Buuren S, Groothuis-Oudshoorn K, van Buuren S, Groothuis-Oudshoorn K. mice: Multivariate Imputation by Chained Equations in R. J. Stat. Softw. 2011;045(i03).

3. van Buuren S et al. mice: Multivariate imputation by chained equations. R package version 2.46.0. 2017.

**Table S1.** **Demographic and Socioeconomic Characteristics in Cases and Controls and Single vs. Recurrent MDD**

| **Outcome** | **Lifetime MDD** | | | | **Recurrent vs Single MDD** | | | |
| --- | --- | --- | --- | --- | --- | --- | --- | --- |
| **Data** | **Training** | | **Test** | | **Training** | | **Test** | |
| **Sample Size** | **642** | **628** | **377** | **3862** | **274** | **273** | **160** | **588** |
| **Group** | **Case** | **Control** | **Case** | **Control** | **Recurrent** | **Single** | **Recurrent** | **Single** |
| **Scottish nationality (N, %)** | 580 (90.3) | 550 (87.6) | 343 (91.0) | 3366 (87.2) | 257 (93.8) | 251 (91.9) | 544 (92.5) | 146 (91.3) |
| **Age (mean, SD)** | 49.6 (11.8) | 56.7 (11.9) | 50.4 (11.7) | 56.9 (11.7) | 47.2 (12.3) | 49.9 (12.4) | 48.7 (12.0) | 51.8 (12.4) |
| **Female (N, %)** | 469 (73.1) | 359 (57.2) | 257 (68.2) | 2162 (56.0) | 202 (73.7) | 195 (71.4) | 417 (70.9) | 106 (66.3) |
| **Living as a couple (N, %)** | 381 (59.3) | 499 (79.5) | 228 (60.5) | 3026 (78.4) | 166 (60.6) | 180 (65.9) | 326 (55.4) | 106 (66.3) |
| **Living alone (N, %)** | 154 (24.0) | 82 (13.1) | 84 (22.3) | 545 (14.1) | 59 (21.5) | 58 (21.2) | 149 (25.3) | 26 (16.3) |
| **Annual personal income** |  |  |  |  |  |  |  |  |
| **£0-£10 000** | 113 (17.6) | 50 (8.0) | 59 (15.6) | 273 (7.1) | 44 (16.1) | 34 (12.5) | 114 (19.4) | 20 (12.5) |
| **£10 001-£30 000** | 243 (37.9) | 181 (28.8) | 127 (33.7) | 1242 (32.2) | 93 (33.9) | 96 (35.2) | 224 (38.1) | 57 (35.6) |
| **£30 001-£50 000** | 126 (19.6) | 169 (26.9) | 95 (25.2) | 987 (25.6) | 65 (23.7) | 76 (27.8) | 115 (19.6) | 32 (20.0) |
| **£50 001 +** | 126 (19.6) | 175 (27.9) | 82 (21.8) | 1002 (25.6) | 59 (21.5) | 54 (19.8) | 114 (19.4) | 38 (23.8) |
| **Refused** | 34 (5.3) | 53 (8.4) | 14 (3.7) | 358 (9.3) | 13 (4.7) | 13 (4.8) | 21 (3.6) | 13 (8.1) |
| **Worked in last 12 months (N, %)** | 393 (61.2) | 420 (66.9) | 242 (64.2) | 2497 (64.7) | 180 (65.7) | 182 (66.7) | 362 (61.6) | 97 (60.6) |
| **Educational Qualification** |  |  |  |  |  |  |  |  |
| **Less than secondary** | 68 (10.6) | 66 (10.5) | 46 (12.2) | 426 (11.0) | 31 (11.3) | 25 (9.2) | 64 (10.9) | 17 (10.6) |
| **Secondary** | 176 (27.4) | 212 (33.8) | 101 (26.8) | 1171 (30.3) | 79 (28.8) | 79 (28.9) | 163 (27.7) | 46 (28.8) |
| **Tertiary** | 398 (62.0) | 350 (55.7) | 230 (61.0) | 2265 (58.6) | 164 (59.9) | 169 (61.9) | 361 (61.4) | 97 (60.6) |
| **SIMD** |  |  |  |  |  |  |  |  |
| **1 - Most deprived** | 111 (17.3) | 60 (9.6) | 70 (18.6) | 401 (10.4) | 65 (23.7) | 37 (13.6) | 107 (18.2) | 18 (11.3) |
| **2** | 112 (17.4) | 73 (11.6) | 63 (16.7) | 440 (11.4) | 40 (14.6) | 36 (13.2) | 111 (18.9) | 23 (14.4) |
| **3** | 116 (18.1) | 107 (17.0) | 53 (14.1) | 15.5 (599) | 43 (15.7) | 48 (17.6) | 98 (16.7) | 32 (20.0) |
| **4** | 144 (22.4) | 148 (23.6) | 87 (23.1) | 1024 (26.5) | 63 (23.0) | 64 (23.4) | 125 (21.3) | 40 (25.0) |
| **5 - Least deprived** | 159 (24.8) | 240 (38.2) | 104 (27.6) | 1398 (36.2) | 63 (23.0 | 88 (32.2) | 147 (25.0) | 47 (29.4) |

Yellow: p<0.005, light green: p<0.01, dark green: p<0.05. Statistical significance was tested between cases/controls for lifetime MDD or between single versus recurrent MDD using χ^2^ tests for categorical and *t*-tests for continuous outcomes.

**Table S2. ROC Curve Comparisons between Methods within Models for Lifetime MDD**

**(a) Model 1 (above diagonal) and Model 2 (below diagonal)**

|  | **C5.0** | **CIF** | **EN** | **FSR** | **GDB** | **NN** | **RF** | **SVM-L** | **SVM-P** | **SVM-R** |
| --- | --- | --- | --- | --- | --- | --- | --- | --- | --- | --- |
| **C5.0** |  | 0.019 | 0.17 | 0.036 | 0.17 | 0.068 | 0.33 | 0.13 | 0.010 | 0.98 |
| **CIF** | 0.029 |  | 0.84 | 0.96 | 0.050 | 0.97 | 0.018 | 0.32 | 0.92 | 0.0066 |
| **EN** | 0.0086 | 0.59 |  | 0.80 | 0.062 | 0.25 | 0.028 | 0.26 | 0.59 | 0.0055 |
| **FSR** | 0.014 | 0.78 | 0.041 |  | 0.80 | 0.43 | 0.037 | 0.12 | 0.87 | 0.012 |
| **GDB** | 0.0086 | 0.51 | 0.20 | 0.81 |  | 0.27 | 0.20 | 0.41 | 0.068 | 0.11 |
| **NN** | 0.065 | 0.94 | 0.54 | 0.61 | 0.55 |  | 0.13 | 0.91 | 0.29 | 0.021 |
| **RF** | 0.020 | 0.91 | 0.36 | 0.81 | 0.58 | 0.86 |  | 0.20 | 0.031 | 0.21 |
| **SVM-L** | 0.17 | 0.46 | 0.46 | 0.0032 | 0.14 | 0.38 | 0.28 |  | 0.31 | 0.030 |
| **SVM-P** | 0.14 | 0.38 | 0.49 | 0.0032 | 0.11 | 0.38 | 0.15 | 0.92 |  | 0.0064 |
| **SVM-R** | 0.94 | 0.024 | 0.067 | 0.0079 | 0.0082 | 0.024 | 0.0073 | 0.12 | 0.043 |  |

**(b) Model 3 (above diagonal) and Model 4 (below diagonal)**

|  | **C5.0** | **CIF** | **EN** | **FSR** | **GDB** | **NN** | **RF** | **SVM-L** | **SVM-P** | **SVM-R** |
| --- | --- | --- | --- | --- | --- | --- | --- | --- | --- | --- |
| **C5.0** |  | 0.12 | 0.0026 | 0.062 | **0.00017** | 0.21 | 0.17 | 0.014 | 0.028 | 0.039 |
| **CIF** | **0.00081** |  | 0.039 | 0.62 | **0.000028** | 0.86 | 0.89 | 0.24 | 0.31 | 0.31 |
| **EN** | **0.0011** | 0.41 |  | 0.26 | 0.37 | 0.080 | 0.035 | 0.41 | 0.072 | 0.15 |
| **FSR** | 0.18 | 0.29 | 0.019 |  | 0.18 | 0.52 | 0.56 | 0.43 | 0.85 | 0.95 |
| **GDB** | **8.65E-08** | **0.00010** | 0.22 | 0.012 |  | 0.012 | 0.0068 | 0.27 | 0.094 | 0.085 |
| **NN** | 0.026 | 0.59 | 0.13 | 0.48 | 0.027 |  | 0.95 | 0.22 | 0.37 | 0.45 |
| **RF** | 0.0089 | 0.69 | 0.22 | 0.37 | 0.014 | 0.75 |  | 0.20 | 0.16 | 0.32 |
| **SVM-L** | 0.0081 | 0.87 | 0.35 | 0.030 | 0.13 | 0.44 | 0.70 |  | 0.54 | 0.45 |
| **SVM-P** | 0.0037 | 0.71 | 0.16 | 0.10 | 0.13 | 0.44 | 0.70 | 0.85 |  | 0.81 |
| **SVM-R** | 0.0082 | 0.94 | 0.15 | 0.13 | 0.054 | 0.49 | 0.66 | 0.88 | 0.52 |  |

*p*-values < 0.05 after Bonferroni correction are in bold.

**Table S3. ROC Curve Comparisons between Methods within Models for Single versus Recurrent MDD**

**(a) Model 1 (above diagonal) and Model 2 (below diagonal)**

|  | **C5.0** | **CIF** | **EN** | **FSR** | **GDB** | **NN** | **RF** | **SVM-L** | **SVM-P** | **SVM-R** |
| --- | --- | --- | --- | --- | --- | --- | --- | --- | --- | --- |
| **C5.0** |  | 0.041 | 0.13 | 0.086 | 0.049 | 0.095 | 0.65 | 0.10 | 0.17 | 0.93 |
| **CIF** | 0.0062 |  | 0.28 | 0.57 | 0.28 | 0.68 | 0.017 | 0.46 | 0.27 | 0.015 |
| **EN** | 0.020 | 0.85 |  | 0.47 | 0.14 | 0.88 | 0.11 | 0.43 | 0.96 | 0.11 |
| **FSR** | 0.19 | 0.016 | 0.071 |  | 0.56 | 0.92 | 0.070 | 0.71 | 0.31 | 0.062 |
| **GDB** | 0.0095 | 0.52 | 0.89 | 0.023 |  | 0.75 | 0.033 | 0.36 | 0.26 | 0.027 |
| **NN** | 0.46 | 0.023 | 0.049 | 0.47 | 0.037 |  | 0.079 | 0.99 | 0.87 | 0.14 |
| **RF** | 0.074 | 0.084 | 0.26 | 0.75 | 0.11 | 0.42 |  | 0.079 | 0.10 | 0.57 |
| **SVM-L** | 0.031 | 0.24 | 0.42 | 0.025 | 0.57 | 0.047 | 0.39 |  | 0.32 | 0.083 |
| **SVM-P** | 0.014 | 0.33 | 0.66 | 0.046 | 0.74 | 0.032 | 0.27 | 0.73 |  | 0.13 |
| **SVM-R** | 0.098 | 0.036 | 0.15 | 0.90 | 0.086 | 0.45 | 0.79 | 0.17 | 0.036 |  |

**(b) Model 3 (above diagonal) and Model 4 (below diagonal)**

|  | **C5.0** | **CIF** | **EN** | **FSR** | **GDB** | **NN** | **RF** | **SVM-L** | **SVM-P** | **SVM-R** |
| --- | --- | --- | --- | --- | --- | --- | --- | --- | --- | --- |
| **C5.0** |  | 0.050 | 0.0052 | 0.67 | **0.00017** | 0.0055 | **0.00019** | 0.78 | 0.36 | 0.47 |
| **CIF** | **1.85E-07** |  | 0.24 | 0.18 | 0.0071 | 0.17 | 0.0053 | 0.10 | 0.36 | 0.25 |
| **EN** | **4.82E-07** | 0.81 |  | 0.0086 | 0.17 | 0.64 | 0.18 | **0.00092** | 0.037 | 0.018 |
| **FSR** | 0.54 | **0.00045** | **0.00012** |  | 0.0022 | 0.020 | 0.0036 | 0.80 | 0.57 | 0.74 |
| **GDB** | **1.62E-06** | 0.54 | 0.72 | **0.00058** |  | 0.58 | 0.68 | **0.00025** | 0.0068 | 0.0023 |
| **NN** | **0.00054** | 0.059 | 0.10 | 0.021 | 0.14 |  | 0.47 | 0.0049 | 0.038 | 0.024 |
| **RF** | **2.26E-07** | 0.88 | 0.74 | **0.00030** | 0.22 | 0.047 |  | **0.00029** | 0.0022 | **0.00038** |
| **SVM-L** | 0.20 | **0.00031** | **4.91E-06** | 0.45 | **0.00024** | 0.032 | 0.032 |  | 0.14 | 0.34 |
| **SVM-P** | 0.014 | 0.020 | 0.023 | 0.077 | 0.054 | 0.47 | 0.014 | 0.054 |  | 0.55 |
| **SVM-R** | 0.024 | 0.0094 | 0.0075 | 0.096 | 0.022 | 0.34 | 0.0043 | 0.054 | 0.52 |  |

*p*-values < 0.05 after Bonferroni correction are in bold.

**Table S4. AUC Differences between MC4 Full Model and Leave-One-Out Analysis of Top 20 Predictors in Lifetime Depression**

| **MC4 Top 20** | **C5.0** | **CIF** | **EN** | **FSR** | **GDB** | **NN** | **RF** | **SVM-L** | **SVM-P** | **SVM-R** |
| --- | --- | --- | --- | --- | --- | --- | --- | --- | --- | --- |
| Neuroticism | 0.034 *(0.00018)* | 0.032 *(3.56e^-05^)* | 0.030 *(2.80e^-08^)* | 0.031 *(1.08e^-07^)* | 0.027 *(1.63e^-05^)* | 0.030 *(2.63e^-07^)* | 0.025 *(6.02e^-06^)* | 0.031 *(4.27e^-08^)* | 0.028 *(2.04e^-07^)* | 0.024 *(3.07e^-05^)* |
| GHQ Total Score | 0.004 | -0.001 | 0.004 | -0.004 | -0.002 | -0.010 | -0.007 | 0.0009 | -0.002 | -0.004 |
| GHQ Depression | -0.004 | 0.0009 | 0.0008 | -0.0002 | 0.003 | -0.010 | -0.004 | 0.005 | -0.006 | 0.0005 |
| GHQ Somatic Symptoms | 0.0009 | -0.003 | 0.004 | -0.002 | 0 | -0.0099 | 0 | 0.002 | -0.002 | -0.0003 |
| Age | 0.008 | 0.003 | 0.009 | 0.0002 | 0.008 | -0.006 | 0.002 | 0.006 | 0.003 | 0.001 |
| Family History: Depression | -0.003 | 0.003 | 0.007 | 0.002 | 0.002 | -0.006 | -0.003 | 0.004 | 0.0008 | -0.0003 |
| Income | 0.001 | -0.002 | 0.003 | -0.004 | -0.001 | -0.0098 | -0.007 | -0.0004 | -0.004 | -0.004 |
| Live as a Couple | 0.008 | 0.0004 | 0.006 | -0.002 | 0.001 | -0.0084 | -0.005 | 0.003 | -0.002 | -0.003 |
| Sex | 0.001 | -0.002 | 0.004 | -0.003 | 0 | -0.010 | -0.007 | 0.002 | -0.001 | -0.003 |
| Mother with Depression | -0.007 | -0.002 | 0.002 | -0.002 | 0 | -0.011 | -0.006 | 0.001 | -0.002 | -0.003 |
| Own House | -0.003 | -0.002 | 0.004 | -0.001 | 0 | -0.0096 | -0.007 | 0.002 | -0.002 | -0.003 |
| GHQ Anxiety | 0.001 | -0.002 | 0.004 | -0.002 | -0.001 | -0.010 | -0.002 | 0.0001 | -0.003 | -0.004 |
| Ever Smoke | 0.0006 | -0.002 | 0.006 | -0.002 | 0.001 | -0.0085 | -0.005 | 0.001 | -0.0001 | -0.002 |
| MDQ Severity Score | -0.002 | 0.0005 | 0.004 | -0.002 | 0 | -0.0087 | -0.004 | 0.002 | -0.002 | -0.002 |
| SPQ Total Score | -0.0004 | -0.003 | 0.005 | -0.002 | -0.001 | -0.011 | -0.007 | -0.002 | -0.0004 | -0.006 |
| Education/Qualifications | 0.005 | 0.002 | 0.006 | -0.002 | 0.002 | -0.0088 | -0.003 | 0.002 | -0.0005 | -0.003 |
| Pain Intensity | -0.0007 | -0.002 | 0.005 | -0.002 | 0 | -0.0093 | -0.007 | 0.001 | -0.002 | -0.003 |
| GHQ Social Dysfunction | -0.003 | -0.002 | 0.004 | -0.002 | -0.001 | -0.010 | -0.0008 | 0.001 | -0.002 | -0.003 |
| Pain Ever | -0.002 | -0.003 | 0.002 | -0.005 | -0.001 | -0.012 | -0.006 | -0.001 | -0.007 | -0.006 |
| Live with People | 0.001 | -0.001 | 0.003 | -0.004 | 0 | -0.010 | -0.007 | 0.0001 | -0.003 | -0.005 |

*p-*values included in parentheses for variables associated with a significant decrease in AUC values after removal, after Bonferroni correction.

**Table S5. AUC Differences between MC4 Full Model and Leave-One-Out Analysis of Top 20 Predictors in Single versus Recurrent Depression**

| **MC4 Top 10** | **C5.0** | **CIF** | **EN** | **FSR** | **GDB** | **NN** | **RF** | **SVM-L** | **SVM-P** | **SVM-R** |
| --- | --- | --- | --- | --- | --- | --- | --- | --- | --- | --- |
| Age at Onset | 0.088 (*0.00056)* | 0.052 | 0.081 (*4.29E^-05^*) | 0.081 (*4.30E^-05^*) | 0.074 (*5.32E^-05^*) | 0.081 (*0.00042*) | 0.068 (*0.00014*) | 0.079 (*2.36E^-05^*) | 0.073 (*5.96E^-05^*) | 0.067 (*0.00032*) |
| Neuroticism | 0.023 | 0.004 | 0.012 | 0.012 | 0.014 | 0.012 | 0.014 | 0.015 | 0.008 | 0.018 |
| GHQ Total Score | 0 | 0 | 0 | 0 | 0.003 | -0.003 | 0 | 0.001 | 0.007 | 0.003 |
| Digit Symbol Substitution | -0.020 | -0.002 | 0 | 0 | -0.002 | -0.008 | -0.004 | 0.001 | -0.009 | -0.006 |
| GHQ Somatic Symptoms | 0.013 | -0.003 | -0.001 | -0.001 | -0.001 | 0 | 0.001 | 0.002 | -0.001 | 0.005 |
| GHQ Depression | -0.006 | 0.001 | -0.001 | -0.001 | 0.003 | -0.005 | 0.001 | 0.001 | -0.004 | -0.001 |
| Age | 0.031 | 0.001 | 0.006 | 0.015 | 0.012 | 0 | 0.005 | 0.005 | 0.002 | 0.007 |
| GHQ Social Dysfunction | -0.008 | 0.001 | -0.001 | -0.001 | 0.006 | -0.009 | 0.006 | 0.001 | 0 | 0.007 |
| Own House | -0.007 | 0 | 0 | 0 | 0 | -0.006 | -0.002 | -0.001 | -0.032 (*0.00047*) | -0.009 |
| Age Started Smoking | -0.001 | 0.004 | 0.003 | 0.003 | 0.004 | -0.009 | 0.004 | 0.005 | -0.005 | 0.006 |

*p-*values included in parentheses for variables associated with a significant decrease in AUC values after removal, after Bonferroni correction.

**Table S6. Effect Size Given by Hedges *g* for Model 3 and MC4 Model in Lifetime and Single versus Recurrent MDD**

|  | **Lifetime MDD: M3 = 154 Variables, MC4 = 20 Variables** | | | | **Recurrent MDD: M3 = 180 Variables, MC4 = 10 Variables** | | | |
| --- | --- | --- | --- | --- | --- | --- | --- | --- |
| **Algorithm** | **M3 *g*** | **M3 *g* 95% CI** | **MC4 *g*** | **MC4 *g* 95% CI** | **M3 *g*** | **M3 *g* 95% CI** | **MC4 *g*** | **MC4 *g* 95% CI** |
| C5.0 | 1.26 | (1.15, 1.37) | 1.23 | (1.12, 1.34) | 0.70 | (0.52, 0.88) | 0.93 | (0.75, 1.12) |
| Conditional inference forest | 1.20 | (1.09, 1.31) | 1.14 | (1.03, 1.25) | 0.87 | (0.69, 1.05) | 0.99 | (0.80, 1.17) |
| Elastic net | 1.41 | (1.30, 1.52) | 1.26 | (1.15, 1.37) | 0.94 | (0.76, 1.12) | 1.13 | (0.95, 1.32) |
| Forward stepwise regression | 1.12 | (1.01, 1.23) | 1.26 | (1.15, 1.37 | 0.62 | (0.44, 0.80) | 1.13 | (0.95, 1.32) |
| Gradient descent boosting | 1.25 | (1.14, 1.35) | 1.23 | (1.12, 1.34) | 1.04 | (0.85, 1.22) | 1.16 | (0.98, 1.34) |
| Neural networks | 1.23 | (1.12, 1.34) | 1.21 | (1.10, 1.32) | 0.97 | (0.78, 1.15) | 1.02 | (0.85, 1.21) |
| Random forest | 1.10 | (0.99, 1.21) | 1.05 | (0.94, 1.16) | 1.02 | (0.84, 1.20) | 1.04 | (0.86, 1.22) |
| SVM, linear | 1.14 | (1.03, 1.22) | 1.16 | (1.06, 1.28) | 0.71 | (0.53, 0.89) | 1.12 | (0.93, 1.30) |
| SVM, polynomial | 1.13 | (1.02, 1.24) | 1.15 | (1.05, 1.26) | 0.80 | (0.62, 0.98) | 0.96 | (0.77, 1.14) |
| SVM, radial basis function | 1.06 | (0.95, 1.17) | 1.12 | (1.01, 1.23) | 0.76 | (0.59, 0.94) | 0.94 | (0.76, 1.12) |
